# The Prevalence of Imposter Phenomenon in (Post)graduate Medical Education

**DOI:** 10.1101/2024.01.07.24300956

**Authors:** Jessica Cheung, Matthew Sibbald, Brandon Ruan, Jonathan Sherbino

## Abstract

**Phenomenon:** Imposter phenomenon (IP) is the feeling of inadequacy despite demonstrating external standards of success. Few studies have broadly examined the prevalence of IP in resident-physicians. This study assessed the prevalence of self-reported IP in resident-physicians, exploring the correlation of demographic risk factors and feelings of IP.

**Approach:** All residents, across all years of training and programs at McMaster University during the 2019-2020 academic year were recruited to complete a self-report survey. Survey items gathered demographic information and measured self-reported feelings of IP and Clance Imposter Phenomenon Scale (CIPS) scores.

**Findings:** 519 out of 977 (53.1%) individuals completed the survey. Measured by the CIPS, clinically significant IP occurred in 59.2% (n=307) of participants. After completing the CIPS, participants self-reported feelings of low (25.0%, n=130), medium (41.9%, n=218), high (19.0%, n=99), and intense (3.7%, n= 19) IP. 64.9% (n=337) of respondents felt they hid feelings of IP during residency. 62.4% (n=324) of respondents were unaware of resources available to them as they struggled with feelings of IP. Only female gender was associated with IP (p <0.001).

**Insights:** IP is highly prevalent across a broad range of residents, independent of clinical discipline and most demographics. Educators and administrators should attend to IP by normalizing the discussion of IP and ensuring adequate access to resources for support.

## Phenomenon

Few studies have broadly examined the prevalence of imposter phenomenon (IP) in resident-physicians. IP is the feeling of inadequacy despite demonstrating external standards of success.^1^ Individuals with feelings of IP often fear exposure of being a fraud and attribute their success to external causes such as charm, luck and hard work, rather than internal causes such as demonstrated intelligence, competence, or ability.^2^ Once a task is assigned, individuals with IP either over-prepare or procrastinate. When the task is accomplished, however, any positive feedback is discounted by an individual with IP. Those who achieved the task through over-preparation attribute success to external causes, such as hard work, rather than internal sources, such as ability. Equally, achievement associated with procrastination is attributed to external causes, such as luck.^2^ Subsequently, both approaches lead to worsening feelings of fraudulence, self-doubt, and increased IP.

Imposter phenomenon has been studied in various groups, including women,^2,3^ undergraduate students, graduate students, nursing students, and business professionals.^1,4^ Studies have shown that individuals experiencing IP are at risk of also experiencing psychological distress from self-oriented perfectionism,^5-7^ anxiety, depression, and low self-confidence.^8-9^

A review of the literature reveals five studies that examined features of IP in attending physicians.^10-14^ Only four small studies, ranging from 48 to 185 participants, have assessed IP in postgraduate medical education: one study of family medicine residents,^15^ two studies of internal medicine residents,^16-17^ and one study of general surgery residents.^14^ There have been no studies to date that assess IP in postgraduate medical education across all specialties, in all years of training.

Recognizing the impact of IP on physician mental health and a lack of research on the prevalence of IP among resident-physicians, the objective of this study was to assess the prevalence of IP in all postgraduate medical education physician-residents at a large Canadian university, and to identify demographic risk factors associated with feelings of IP.

## Approach

An online survey was administered to all residents enrolled in a McMaster University residency training program between March 3 and 10, 2020. Thus, the recruitment period began on March 3, 2020 and concluded on March 10, 2020. Participation in this between-subjects study was voluntary and written informed consent was obtained through email. There were no minors recruited in this study.

The survey consisted of questions on participant demographics, self-perceived IP, available resources to combat IP, and self-rating on the Clance Imposter Phenomenon Scale (CIPS). CIPS is a research standard, peer reviewed instrument to measure IP.^18-19^ CIPS consists of 20 questions on a 5-point Likert scale from “not at all true” to “very true.” The questions self-assess agreement with statements regarding task completion and performance evaluation. No modifications were made to the instrument. Total scores range from 20 to 100. A total score of 40 or less indicates few IP experiences, a score between 41 and 60 indicating moderate IP experiences, a score of 61 to 80 indicating frequent IP experiences, and a score of 81 to 100 indicating intense IP experiences. Clance established a score of 62 or greater to indicate clinically significant IP, meeting criteria to be diagnosed as an imposter.^19^ This cut point in the original research on IP ensures no false negatives and minimal false positive determinations of IP. It is the current standard for using CIPS. Self-perceived IP was measured to determine if a simpler instrument could replace CIPS.

Two faculty members and two research assistants piloted tested the survey for clarity and usability. They subsequently participated in cognitive interviews conducted by the senior investigator, who has experience in survey design, to identify necessary modifications to the final version of the survey instrument. See the supplementary appendix (S1-S2) for the survey instrument. A link to the survey was emailed once a week for a total of four weeks. All survey responses were anonymized; data was unavailable to program directors and administrators.

Survey data was reported using descriptive statistics. Univariate analysis of variance was performed to compare CIPS scores across categorical variables. Accepting a 5% probability of type I error, the threshold for statistical significance was set at a p<0.006 based on a Bonferonni correction for multiple comparisons.^20^ Self-report of IP and CIPS scores were compared using a weighted kappa given the ordinal nature of both variables. Statistics were performed using SPSS Software version 26 (IBM, Redmond). Free text responses were aggregated into conceptual categories.^21^

The study was approved by the Hamilton Integrated Research Ethics Board (protocol number: 4597).

## Findings

Of the 977 residents invited to participate in the study, 519 completed the survey (53.1%). Table 1 reports participant demographics.

**Table 1:**
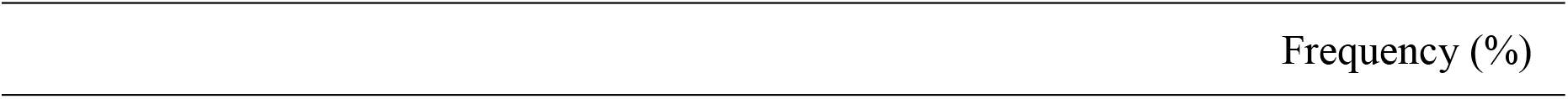
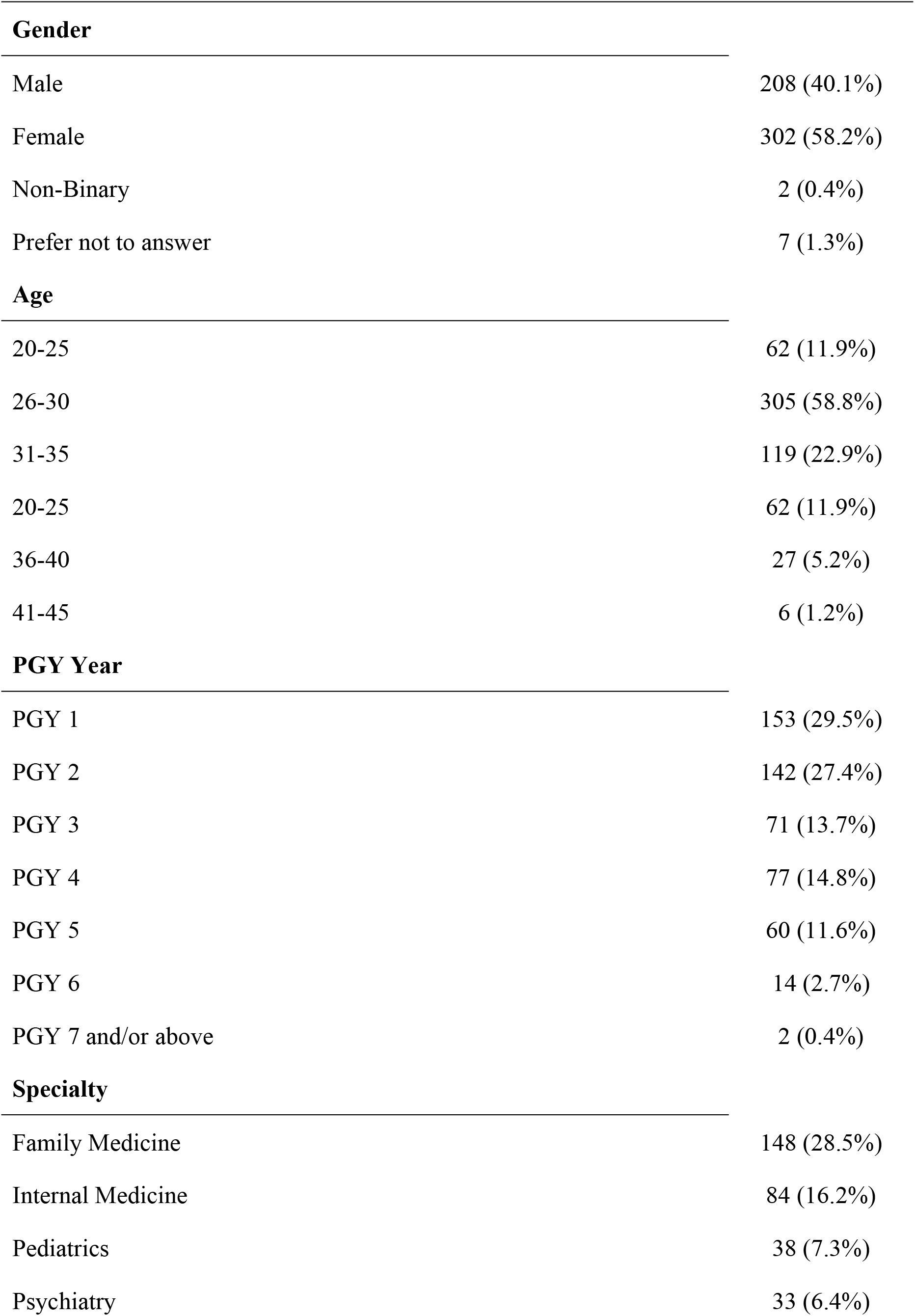
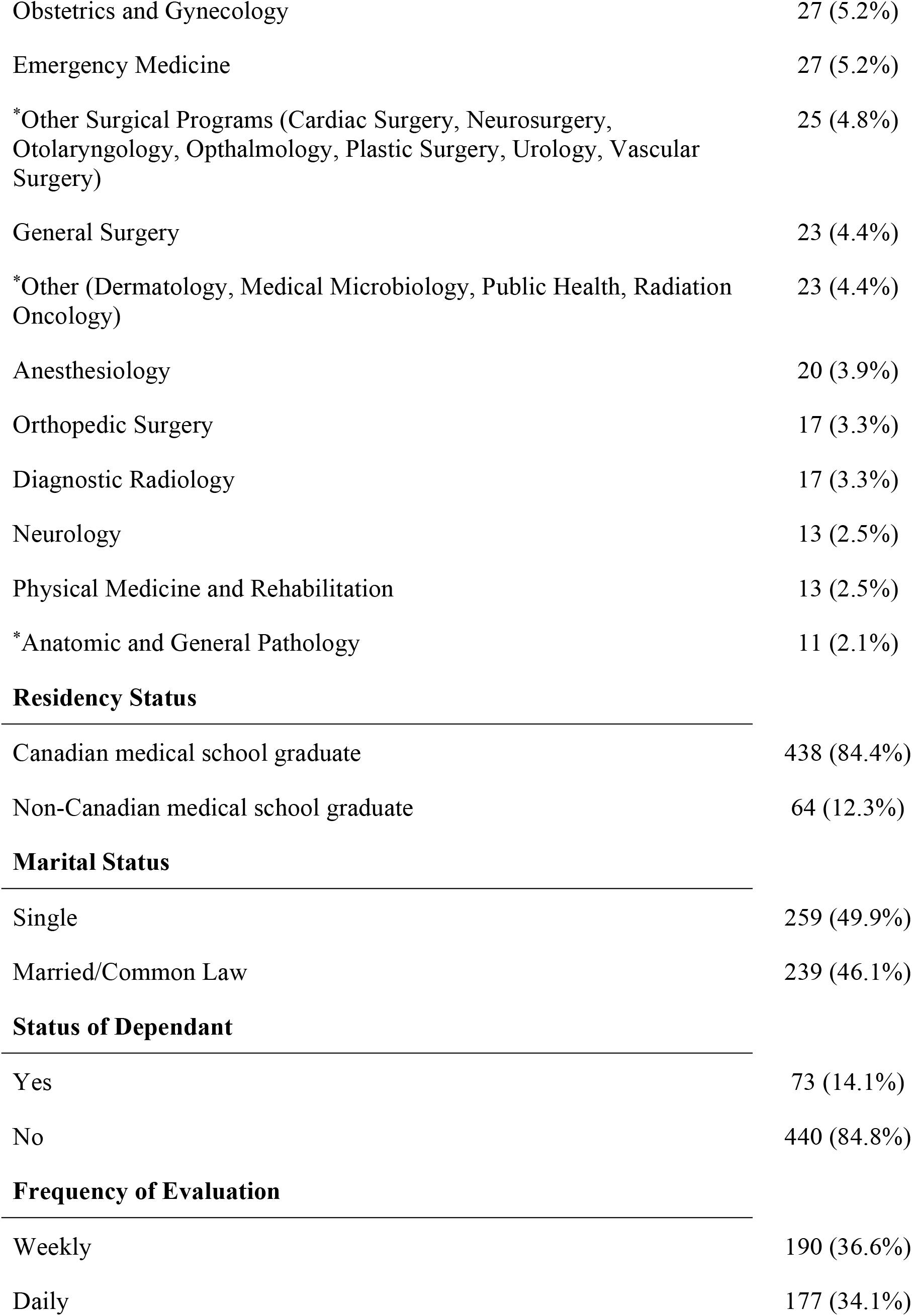
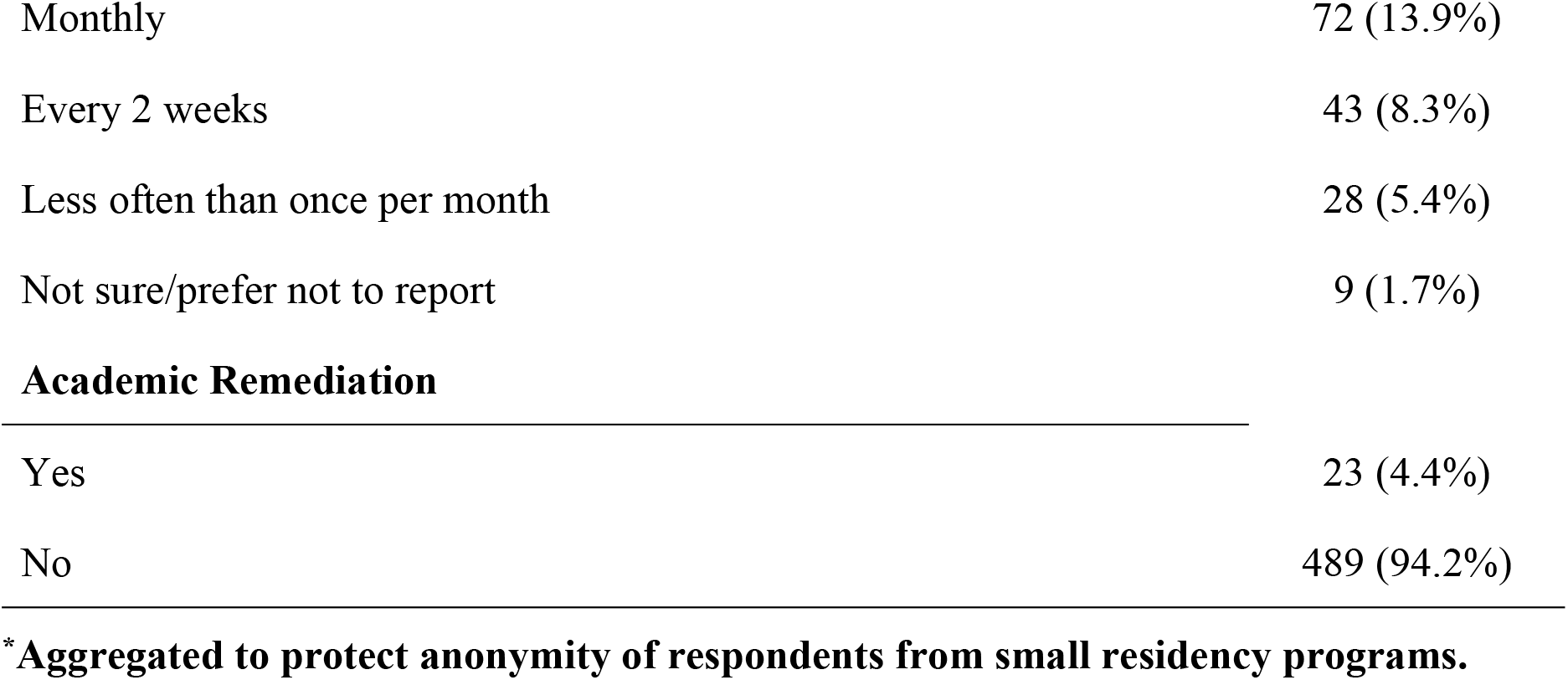
Participant Demographics.

|  | Frequency (%) |
| --- | --- |
| **Gender** |  |
| Male | 208 (40.1%) |
| Female | 302 (58.2%) |
| Non-Binary | 2 (0.4%) |
| Prefer not to answer | 7 (1.3%) |
| **Age** |  |
| 20-25 | 62 (11.9%) |
| 26-30 | 305 (58.8%) |
| 31-35 | 119 (22.9%) |
| 20-25 | 62 (11.9%) |
| 36-40 | 27 (5.2%) |
| 41-45 | 6 (1.2%) |
| **PGY Year** |  |
| PGY 1 | 153 (29.5%) |
| PGY 2 | 142 (27.4%) |
| PGY 3 | 71 (13.7%) |
| PGY 4 | 77 (14.8%) |
| PGY 5 | 60 (11.6%) |
| PGY 6 | 14 (2.7%) |
| PGY 7 and/or above | 2 (0.4%) |
| **Specialty** |  |
| Family Medicine | 148 (28.5%) |
| Internal Medicine | 84 (16.2%) |
| Pediatrics | 38 (7.3%) |
| Psychiatry | 33 (6.4%) |
| Obstetrics and Gynecology | 27 (5.2%) |
| Emergency Medicine | 27 (5.2%) |
| *Other Surgical Programs (Cardiac Surgery, Neurosurgery, Otolaryngology, Ophthalmology, Plastic Surgery, Urology, Vascular Surgery) | 25 (4.8%) |
| General Surgery | 23 (4.4%) |
| *Other (Dermatology, Medical Microbiology, Public Health, Radiation Oncology) | 23 (4.4%) |
| Anesthesiology | 20 (3.9%) |
| Orthopedic Surgery | 17 (3.3%) |
| Diagnostic Radiology | 17 (3.3%) |
| Neurology | 13 (2.5%) |
| Physical Medicine and Rehabilitation | 13 (2.5%) |
| *Anatomic and General Pathology | 11 (2.1%) |
| **Residency Status** |  |
| Canadian medical school graduate | 438 (84.4%) |
| Non-Canadian medical school graduate | 64 (12.3%) |
| **Marital Status** |  |
| Single | 259 (49.9%) |
| Married/Common Law | 239 (46.1%) |
| **Status of Dependant** |  |
| Yes | 73 (14.1%) |
| No | 440 (84.8%) |
| **Frequency of Evaluation** |  |
| Weekly | 190 (36.6%) |
| Daily | 177 (34.1%) |
| Monthly | 72 (13.9%) |
| Every 2 weeks | 43 (8.3%) |
| Less often than once per month | 28 (5.4%) |
| Not sure/prefer not to report | 9 (1.7%) |
| Academic Remediation |  |
| Yes | 23 (4.4%) |
| No | 489 (94.2%) |

\*Aggregated to protect anonymity of respondents from small residency programs.

Clinically significant IP, as measured by a CIPS score greater than 61, occurred in 59.2% (n=307) of participants. CIPS scores of high (score of 60-79) or intense (80-100) IP occurred in 46.8% (n=243) and 14.6% (n=76) participants, respectively. Participants self-report of whether they have IP and their degree of IP demonstrated low (25.0%, n=130), medium (41.9%, n=218), high (19.0%, n=99), and intense (3.7%, n= 19) IP. 10.4% of participants (n = 54) reported no feelings of IP. Self-reported feelings of IP correlated moderately with CIPS (weighted kappa = 0.58).

Female gender was the only demographic factor that contributed to variance in CIPS scores. Participants identifying as female (n=302) had a mean CIPS score of 66, compared to mean CIPS score of 61 in respondents who identified as male (n=208) and a mean CIPS score of 59 in respondents who identified as nonbinary/preferred not to report (n=9) (p <0.001). Older age, marital status, being a parent, postgraduate year training level, clinical specialty, international medical graduate status, frequency of assessments in residency, and previous academic remediation did not predict IP. (See the supplementary appendix, S3, for all comparisons.)

Approximately two-thirds (64.9%, n=337) of respondents hid their feelings of inadequacy during residency. Similarly, nearly two-thirds (62.4%, n=324) of respondents were previously unaware of resources to help them address IP. Table 2 includes resources that participants identified as potentially helpful for addressing IP.

**Table 2:**
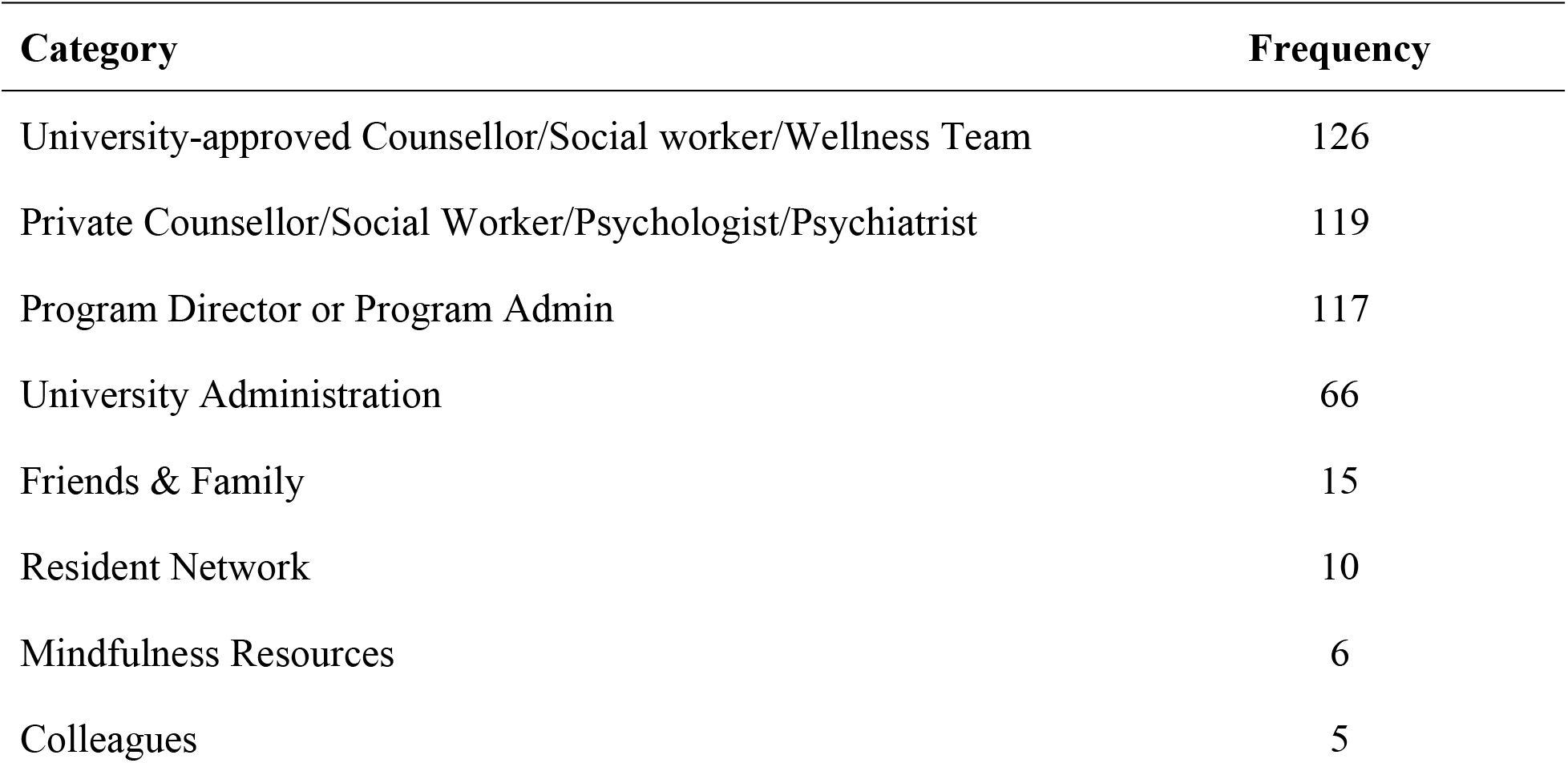
List of support networks that residents consult for feelings of imposter phenomenon.

| Category | Frequency |
| --- | --- |
| University-approved Counsellor/Social worker/Wellness Team | 126 |
| Private Counsellor/Social Worker/Psychologist/Psychiatrist | 119 |
| Program Director or Program Admin | 117 |
| University Administration | 66 |
| Friends & Family | 15 |
| Resident Network | 10 |
| Mindfulness Resources | 6 |
| Colleagues | 5 |

## Insights

This study suggests a high prevalence of imposter phenomenon in residents, across a diversity of clinical disciplines. A majority of participants (59.2%) met the standard of clinically significant IP as measured by the CIPS.

This study demonstrates a greater pervasiveness of IP in residents across a diversity of disciplines as compared to smaller, single-specialty studies (33% in family medicine residents^15^ and approximately 40% in internal medicine residents.^16^) The majority of respondents hid their feelings of inadequacy. Fear of discrimination, remediation, impact on academic and employment opportunities may play a role in preventing a resident from identifying their IP and seeking formal support from educators and program administrators. Most respondents were unaware of available resources.

Educators and administrators should attend to IP by normalizing the discussion of IP and ensuring adequate access to resources for support. Several studies have looked at successful implementation of low-cost resiliency curricula, which may help address root causes of IP.^22-23^ Resiliency curricula have incorporated strategies including meditation, behavioral skills, setting realistic goals, managing expectations, and gratitude exercises in both didactic and group-based delivery.^24-26^ A systematic review identified resiliency workshops and cognitive behavioral training as the strongest interventions for developing healthcare worker resilience.^27^ The impact of all of these interventions at ameliorating physician IP is modest. The study by Gottlieb et al., found that social supports, validation of success, and personal as well as shared reflections were protective against IP.^4^ Education curricula that include peer-to-peer support, endorse sharing of failures and celebrating success may ameliorate the impact of IP on individuals.

An alternative interpretation of the high prevalence of IP is that signals a fundamental dysfunction of residency education. The rate of burnout in residents has remained unchanged over the past two decades despite substantive world-wide increases in policies and educational interventions targeting resident needs.^28^ Like burnout, IP may be correlated to the system of education, where interventions targeting the resident may be inefficient or ineffective.^29^ Future research should address system changes to ameliorate IP, rather than focusing exclusively on the learner.

This study suggests a difference in CIPS stratified by gender. While females had a mean CIP score consistent with clinically significant IP, males had a mean CIP score just below the cutoff for clinically significant IP. A systematic review by Bravata et al. examined IP and gender.^3^ Sixteen studies found IP to be higher in females than in males, while 17 studies found no difference. A review by Gottlieb et al.^4^, focused on physicians, found 5 studies where IP was more prevalent in women in both undergraduate^5, 30,31^ and postgraduate training,^15,16^ while a similar number of studies found no gender differences.^14,17, 32-34^ Within medicine, there remains significant gender inequality in leadership roles, remuneration, and career trajectories.^35^ Tackling these inequities represents a potential systematic approach to reducing IP which has been highlighted in recent reviews. Suggested interventions include: tackling persistent institutional stereotypes, educational interventions to address microaggressions, increasing diversity via recruitment and enacting systematic process for equitable treatment of all groups.^36,37^ While contributing elements are multifactorial and systems are complex, future research about IP should assess the association of IP, gender, and other inequities in medicine.

Participants were asked “do you think you have features of imposter phenomenon?” and “ … how strong are your feelings…?” The prevalence of self-perceived IP was similar to CIPS scores. However, self-determination of IP is insufficiently correlated to the 20-question CIPS to justify adoption of a simpler instrument.

As a Canadian, single-center study, the generalizability of these findings to other centers or jurisdictions is limited. However, it is the largest and only multi-specialty resident study to date, improving on previous much smaller and discipline-specific studies. The study is also limited by a design that relies on self-reported data. However, this is a feature notable in all the IP literature. Women and non-surgical specialties are over-represented among respondents. Non-responders may reflect different perceptions about IP. In theory, non-responders could counter the strong signal from participants, mitigating the study conclusions. While self-report data is readily influenced by social desirability bias, we believe the strong endorsement for a negative attribute suggests this may not be a substantive limitation of this study.

## Conclusions

In a large, single-centre study, there is a high prevalence of imposter phenomenon in residents, independent of clinical discipline and most demographic factors. Educators and administrators should normalize the discussion of IP and provide resources so that residents and residency programs can address feelings of inadequacy.

## Data Availability

All relevant data are within the manuscript and its Supporting Information files.

## Acknowledgements

The authors thank Drs. Mark Crowther (Chair, Department of Medicine, McMaster University) and Parveen Wasi (Associate Dean, Postgraduate Medical Education, McMaster University) for support in completing this study. The authors also thank Heba Khan (MERIT research assistant) for help in manuscript preparation.

## Ethical Approval

This study was approved by the Hamilton Integrated Research Ethics Board (protocol number: 4597).

